# Concurrent validity and discriminative ability of force plate measures of balance during the sub-acute stage of stroke recovery

**DOI:** 10.1101/2024.04.18.24306027

**Authors:** Raabeae Aryan, Kara K. Patterson, Elizabeth L. Inness, George Mochizuki, Avril Mansfield

## Abstract

**Background:** Many objective measures of balance control, including force plate measures of standing balance, lack sufficient validation for use in the stroke population.

**Research questions:** Do force plate measures of quiet standing balance during the sub-acute stage of stroke recovery have concurrent validity (i.e., correlate with functional balance measures) and discriminative ability (i.e., differentiate fallers from non-fallers and/or those with low-moderate versus high risk of falling)?

**Methods:** Participants completed one trial of quiet standing with eyes open, lasting for 30 seconds. Mean speeds of centre of pressure along the anterior-posterior and medial-lateral axes, weight-bearing asymmetry, and symmetry index were calculated. Concurrent validity of these measures were established against the Berg Balance Scale; their abilities in differentiating fallers from non-fallers, and individuals with low-moderate versus high risk of falling were evaluated using the area under the receiver operating curve (AUC).

**Results:** Among the measures studied, mean speed of centre of pressure along the anterior-posterior axis demonstrated the strongest correlation with the Berg Balance Scale (ρ=-0.430, p-value=0.01). Weight-bearing asymmetry showed the highest ability in differentiating fallers from non-fallers (AUC= 0.69), as well as individuals with low-moderate versus high risk of falling (AUC= 0.66).

**Significance:** Our findings suggest that speed of centre of pressure along the anterior-posterior axis, and weight-bearing asymmetry are valid for use in the sub-acute stage of stroke recovery. These validated measures can better inform rehabilitation practice about the ability of upright standing balance following a stroke.

## 1. INTRODUCTION

Debilitating stroke sequelae, such as spasticity, hemiparesis, balance impairments, and falls,^1,2^ necessitate comprehensive evaluations of postural balance using validated measures. While force plate measures are commonly used in balance research, and can offer useful information about the mechanisms of balance deficits,^3,4^ their validation for post-stroke use remains unclear.

Validity of a measure implies its ability to accurately measure what it is intended to measure, and informs the interpretability of and conclusions drawn from its results.^5^ Thus, it is imperative to validate measurement tools in the context of populations of interest,^5^ such as for use after a stroke.

Although some force plate measures have been validated in various non-stroke populations,^6,7^ and some prior studies investigated the relationships among the performance-based measures (e.g., Tinetti test, Timed Up and Go, Berg Balance Scale) and some force plate measures (e.g., weight-bearing asymmetry, CoP velocity, ellipse area) in stroke,^8–11^ validation of commonly used force plate measures of quiet upright standing has not been explicitly specified as a research objective in studies within the sub-acute stage of stroke recovery. The previous work in stroke has been conducted either in chronic stroke,^9–11^ or on a mixed sample of individuals across diverse recovery stages (from sub-acute to chronic),^8^ but not entirely within the sub-acute stage of stroke recovery. This is important, because the rate of neurological and functional changes and improvements during the sub-acute stage (i.e., more than 7 days to 6 months post-stroke),^12^ particularly during the first 3 months, is much greater than the chronic stage of stroke recovery.^13,14^

We previously determined that mean speeds of centre of pressure (CoP) along the anterior-posterior (AP) and medial-lateral (ML) directions, weight-bearing asymmetry (WBA), and symmetry index had high test-retest reliability (ICC≥0.82) and low measurement error among people with sub-acute stroke.^15^ Furthermore, reportedly these measures are clinically useful as they can represent the neuromuscular control generated mainly by the muscles of the lower limbs to regulate postural sway (speeds of CoP), quantify asymmetry in distribution of body-weight on the paretic and non-paretic sides (WBA), and address individual-limb contribution to balance control (symmetry index) respectively.^16–18^

Therefore, in this work we aimed to validate these force plate measures of quiet standing balance during the sub-acute stage of stroke recovery. To this end, our primary objectives were to determine the 1) concurrent validity; and 2) ability of this set of measures in classifying individuals with stroke into sub-groups with and without a history of falls, or into sub-groups with various risks of falling. Our secondary objective was to determine the correlations between the same force plate measures of standing balance and measures of motor impairment of the paretic limb,^19^ and balance confidence.^20^

## 2. METHODS

### 2.1. Participants

The force plate time-series processed in this study were extracted from a dataset created based on a larger observational study,^3^ in which participants were recruited between September 2010 and October 2013. Eligible individuals for that original study were people within the sub-acute stage post-stroke, who additionally were: a) admitted to the Toronto Rehabilitation Institute for post-stroke inpatient rehabilitation, b) 18 years of age and older, c) able to stand independently for at least one minute, and d) able to understand test instructions in English. Individuals with other neurologic/orthopedic pathologies, potentially impacting their balance control, were ineligible. The original study was approved by the Research Ethics Board of the University Health Network, Toronto, Canada. Participants provided written informed consent.

Participants were eligible for the current study if they had a 30-second force plate-based assessment of standing balance at discharge and: 1) a Berg Balance Scale assessment (BBS) score obtained within 3 days of their force plate assessment; 2) information about occurrence of falls during the acute-care stay; and/or 3) risk of falling at admission to inpatient rehabilitation, evaluated by the St. Thomas Risk Assessment Tool in Falling Elderly Inpatients (STRATIFY). Demographic and clinical characteristics of eligible participants were either obtained directly from the participants or extracted from their medical records.

### 2.2. Quiet standing assessment and processing

Participants stood on two side-by-side force plates (25*50 cm each, ∼0.1 cm between-plates gap; OR6-7-2000, Advanced Medical Technology Inc., Watertown, Massachusetts, USA) with eyes open and with each foot placed on one force plate in a standard position (toe out: 14 degrees; heels centre-to-centre: 17 cm).^21^ Participants took rests as needed during the sessions; to ensure their safety, participants were supervised by a physiotherapist and a research assistant.

Force plate data were sampled at 256 Hz, and filtered at 10 Hz using a zero phase-lag low-pass 4^th^-order Butterworth filter offline. Then, CoP time-series along the AP and ML-directions (under both feet combined (net-CoP), and under each foot separately) were calculated, down-sampled to 64 Hz, and detrended. The following measures were calculated via custom-written MATLAB codes (2016b, The MathWorks Inc., Natick, Massachusetts, USA):

a. Mean speeds of CoP were calculated by dividing the net-CoP path length, separately along the AP and ML-directions, by the test duration.^22^
b. WBA was the mean vertical ground reaction force under the paretic limb, expressed as the percentage of the total mean vertical ground reaction forces under both feet combined.^16^
c. Symmetry index was calculated by dividing the root mean square (RMS) of the speed of AP-CoP of the non-paretic side by the sum of the RMS of speed of AP-CoP of both non-paretic and paretic sides.^15^

### 2.3. Clinical assessments

Performance-based balance ability, self-perceived balance confidence, and the level of paretic limb motor impairment were evaluated respectively using the BBS,^23^ Activities-specific Balance Confidence scale^20^ (ABC; ICC=0.85 for test-retest reliability in stroke),^24^ and the Chedoke-McMaster Stroke Assessment^19^ (CMSA; ICCs=0.98 and 0.85 respectively for intra-rater and inter-rater reliabilities of leg score, and ICCs=0.94 and 0.96 for intra-rater and inter-rater reliabilities of foot score, both in stroke).^19^ We investigated concurrent validity of force plate balance measures against the BBS, which is among the most widely used performance-based balance measures.^25^ The BBS has high reliability for use in stroke,^26^ and has previously shown a significantly high correlation with the mean speed of CoP in people with chronic stroke (r=-0.760).^9^ These scales were administrated by the participants” physiotherapists as parts of their routine care, and their results were extracted from the hospital charts. When multiple ABC and CMSA records were available, those performed closer to the BBS date were selected.

#### History of falls

Information about occurrence of falling in acute-care was extracted from the hospital charts, incident reports, and participant recall. Accordingly, we separated individuals into “fallers” (those with at least one fall incident) and “non-fallers” (no fall during acute-care stay).

#### Risk of falling

Information about risk of falling was determined by using the STRATIFY score; this score has shown to have a sensitivity of 0.35 but a high specificity of 0.93 in predicting patients at risk of falling.^27^ The STRATIFY classifies individuals into those at low, moderate, and high risk of falling; however, in our study we combined those with low and moderate risk into one group, and those with high risk formed the other group.

### 2.4. Statistical analysis

Normality assumptions were investigated using Shapiro-Wilks” normality test. Concurrent validity of force plate measures, and their correlations with ABC and CMSA were studied using the Spearman”s rank correlation coefficients. Correlation coefficients≥0.4 were considered as suggesting moderate or greater concurrent validity.^5,28,29^

The ability of force plate balance measures in discriminating fallers from non-fallers, or those with low-moderate risks from individuals at high risk of falling were studied using the area under (AUC) the receiver operating characteristics (ROC) curve. We considered measures with AUC values≤0.5 as unable to differentiate, 0.6-0.7 as poor discriminators, 0.7-0.8 as fair discriminators, 0.8-0.9 as good discriminators, and ≥0.9 as excellent discriminators.^30,31^ The Youden method was used to determine the cut-off values (the point closest to the top left corner of the ROC curve) for those force plate measures that achieved an AUC>0.6.^32^ Statistical significance level was set as α=0.05. Statistical analysis was completed using R software (v3.3.2; R Core Team, 2017).

## 3. RESULTS

Data from 33 out of 104 original participants were included in concurrent validity and correlational analyses, and data from 51/104 participants were included in the analysis of discriminate ability. Demographic characteristics are presented in Table 1; descriptive statistics of force plate measures for both the concurrent validity and discriminative ability analyses are presented in Table 2. Fourteen out of 51 participants (27.5%) had at least one fall during acute-care stay, and 19 out of 51 had high risk of falling (37.3%). Except for one participant in the concurrent validity, and one in discriminative ability analyses (tested 102, and 92 days post-stroke, respectively), all participants were assessed during the early sub-acute stage (first 3 months post-stroke).

**Table 1:**
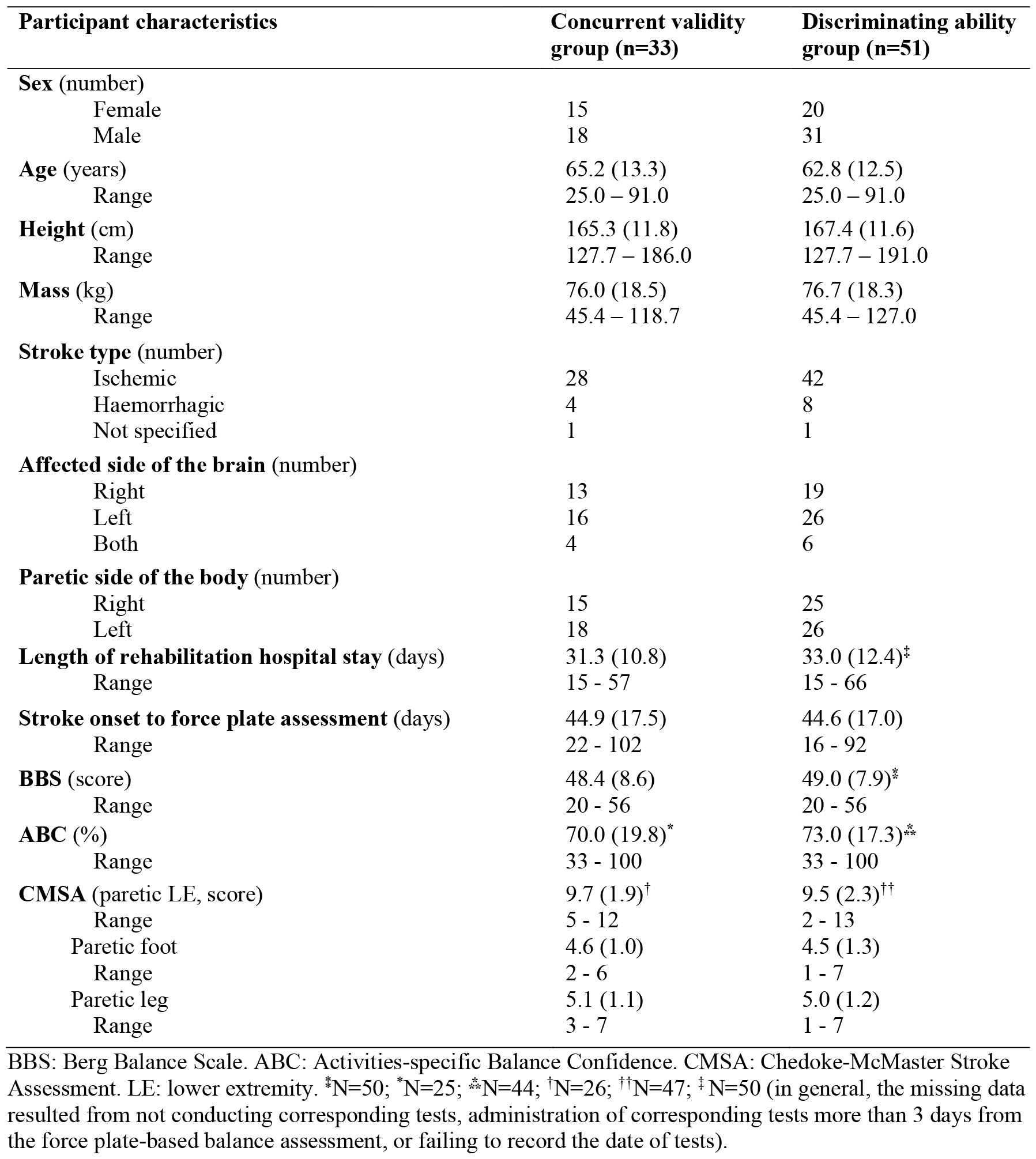
Participants characteristics. Values presented are means, with standard deviations in parentheses for continuous variables and counts for categorical variables.

| Participant characteristics | Concurrent validity group (n=33) | Discriminating ability group (n=51) |
| --- | --- | --- |
| **Sex (number)** |  |  |
| Female | 15 | 20 |
| Male | 18 | 31 |
| **Age (years)** | 65.2 (13.3) | 62.8 (12.5) |
| Range | 25.0 – 91.0 | 25.0 – 91.0 |
| **Height (cm)** | 165.3 (11.8) | 167.4 (11.6) |
| Range | 127.7 – 186.0 | 127.7 – 191.0 |
| **Mass (kg)** | 76.0 (18.5) | 76.7 (18.3) |
| Range | 45.4 – 118.7 | 45.4 – 127.0 |
| **Stroke type (number)** |  |  |
| Ischemic | 28 | 42 |
| Haemorrhagic | 4 | 8 |
| Not specified | 1 | 1 |
| **Affected side of the brain (number)** |  |  |
| Right | 13 | 19 |
| Left | 16 | 26 |
| Both | 4 | 6 |
| **Paretic side of the body (number)** |  |  |
| Right | 15 | 25 |
| Left | 18 | 26 |
| **Length of rehabilitation hospital stay (days)** | 31.3 (10.8) | 33.0 (12.4)<sup>‡</sup> |
| Range | 15 - 57 | 15 - 66 |
| **Stroke onset to force plate assessment (days)** | 44.9 (17.5) | 44.6 (17.0) |
| Range | 22 - 102 | 16 - 92 |
| **BBS (score)** | 48.4 (8.6) | 49.0 (7.9)<sup>‡</sup> |
| Range | 20 - 56 | 20 - 56 |
| **ABC (%)** | 70.0 (19.8)<sup>*</sup> | 73.0 (17.3)<sup>**</sup> |
| Range | 33 - 100 | 33 - 100 |
| **CMSA (paretic LE, score)** | 9.7 (1.9)<sup>†</sup> | 9.5 (2.3)<sup>††</sup> |
| Range | 5 - 12 | 2 - 13 |
| Paretic foot | 4.6 (1.0) | 4.5 (1.3) |
| Range | 2 - 6 | 1 - 7 |
| Paretic leg | 5.1 (1.1) | 5.0 (1.2) |
| Range | 3 - 7 | 1 - 7 |

BBS: Berg Balance Scale. ABC: Activities-specific Balance Confidence. CMSA: Chedoke-McMaster Stroke Assessment. LE: lower extremity. <sup>\*</sup>N=50; <sup>\*</sup>N=25; <sup>\*\*</sup>N=44; <sup>†</sup>N=26; <sup>††</sup>N=47; <sup>‡</sup>N=50 (in general, the missing data resulted from not conducting corresponding tests, administration of corresponding tests more than 3 days from the force plate-based balance assessment, or failing to record the date of tests).

**Table 2:** Descriptive statistics of force plate measures of standing balance for groups in the concurrent and discriminating ability analyses. Values presented are means, with standard deviations in parentheses.

| Force plate measures | Concurrent validity group (n=33) | Discriminating ability group (n=51) |
| --- | --- | --- |
| Mean Speed AP-CoP (mm/s) | 11.9 (5.6) | 11.2 (4.9) |
| Mean Speed ML-CoP (mm/s) | 6.0 (2.9) | 5.8 (2.8) |
| WBA (%body weight) | 46.5 (9.6) | 46.8 (8.6) |
| Symmetry index | 0.60 (0.14) | 0.60 (0.15) |

AP: anterior-posterior. ML: medial-lateral. CoP: net-centre of pressure. WBA: weight-bearing asymmetry.

Fig. 1A shows the strength of concurrent validity of force plate balance measures, plotted against the BBS. Mean speed of AP-CoP demonstrated a statistically significant moderate and negative correlation with the BBS (ρ=-0.430, p-value=0.01). Mean speed of ML-CoP showed a trend towards a weak negative correlation with the BBS, although not statistically significant (ρ=-0.310, p-value=0.08). Negligible correlations were observed between the other two force plate measures and the BBS (p-values>0.76).

**Fig. 1:**
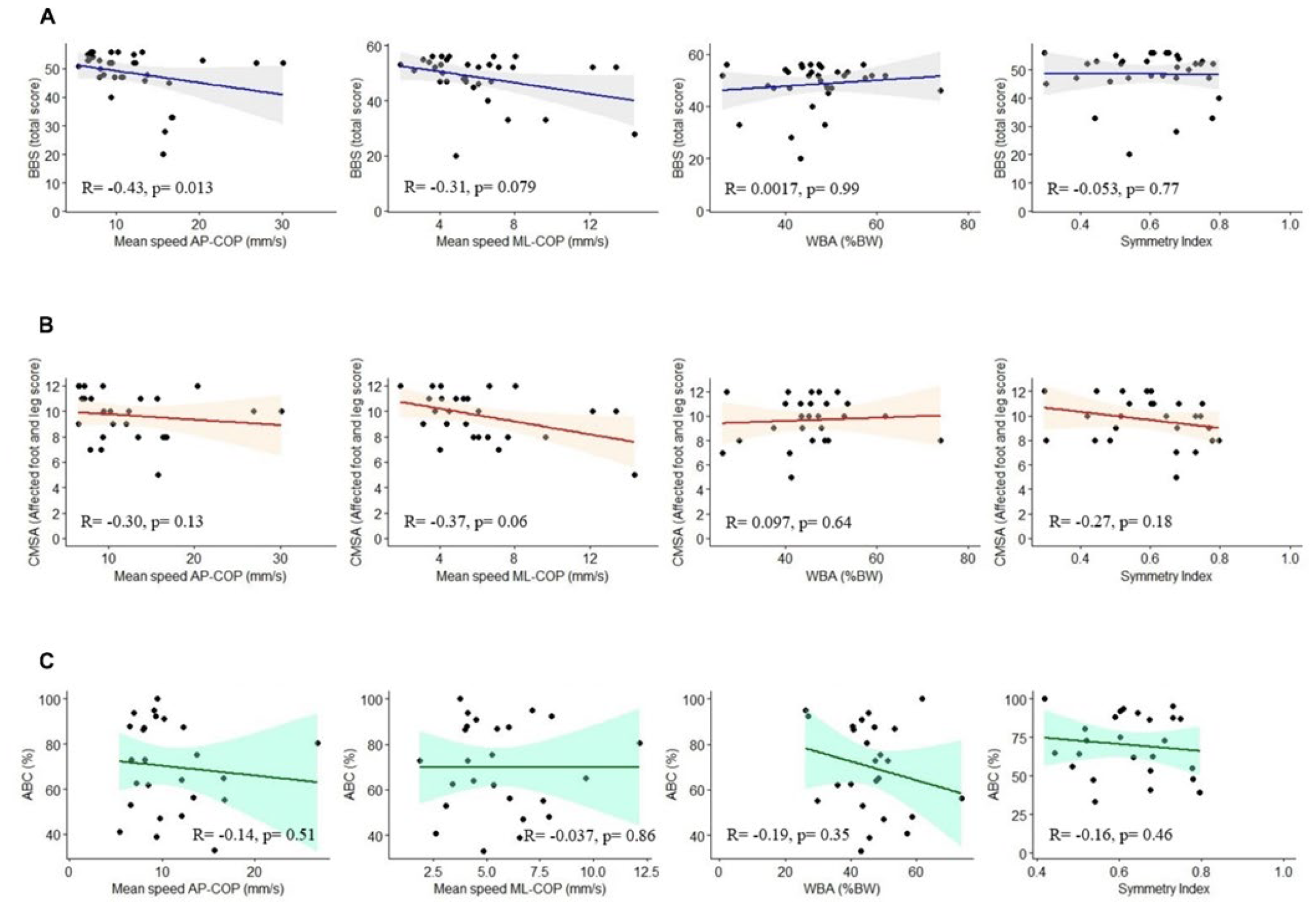
**(A)** Relationships between the force plate measures of standing balance (x-axis) and Berg Balance Scale (BBS) in sub-acute stage post-stroke. **(B)** Scatter plots of correlations between the force plate measures and Activities-specific Balance Confidence (ABS), and **(C)** Chedoke-McMaster Stroke Assessment (CMSA)-paretic foot and leg score. WBA: weight-bearing asymmetry. %BW: percentage of body weight on paretic side. COP: net-centre of pressure. AP: anterior-posterior. ML: medial-lateral.

Scatter plots in Fig. 1B and 1C demonstrate the correlations between the force plate measures of standing balance and the ABC and CMSA-foot and leg scores. Although our results show no statistically significant correlations between the force plate measures and the ABC or CMSA-foot and leg scores, mean speed of ML-COP and CMSA showed the highest correlation overall (p-value=0.06).

The ability of force plate measures of standing balance in differentiating known sub-groups of individuals within the sub-acute stage of stroke recovery are presented in Table 3. Among all force plate measures studied here, WBA demonstrated the highest ability in differentiating fallers from non-fallers (AUC=0.69, CI_95_%=[0.53, 0.86]). Using the Youden method, the cut-off value for the WBA was 47.8% of body weight on the paretic side; this threshold has a sensitivity of 85.7% and specificity of 56.8% in detecting fallers with paretic weight-bearing below the cut-off value. The ability of the other force plate measures to discriminate between fallers and non-fallers was negligible (0.51<AUC<0.56).

**Table 3:**
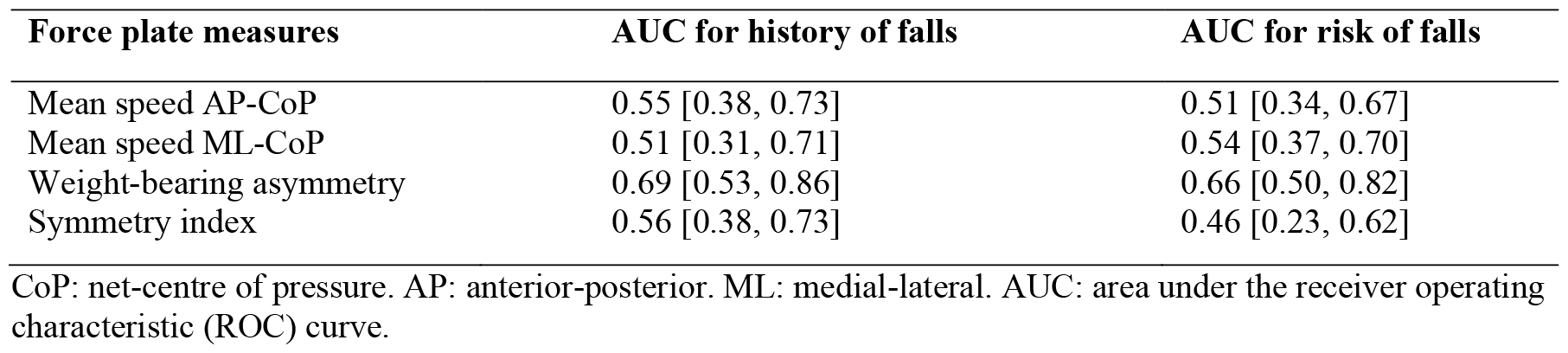
Ability of the force plate measures to discriminate between participants with and without a history of falls, and between participants at high versus low-moderate risk of falls. Values presented are areas under the ROC curve with 95% confidence intervals in brackets.

| Force plate measures | AUC for history of falls | AUC for risk of falls |
| --- | --- | --- |
| Mean speed AP-CoP | 0.55 [0.38, 0.73] | 0.51 [0.34, 0.67] |
| Mean speed ML-CoP | 0.51 [0.31, 0.71] | 0.54 [0.37, 0.70] |
| Weight-bearing asymmetry | 0.69 [0.53, 0.86] | 0.66 [0.50, 0.82] |
| Symmetry index | 0.56 [0.38, 0.73] | 0.46 [0.23, 0.62] |

CoP: net-centre of pressure. AP: anterior-posterior. ML: medial-lateral. AUC: area under the receiver operating characteristic (ROC) curve.

Similarly, WBA showed the highest ability in differentiating participants with low-moderate versus high risk of falling (AUC=0.66, CI_95_%=[0.50, 0.82]). The cut-off value for WBA was 46.2% of body weight on the paretic side; this threshold shows a sensitivity of 68.4% and specificity of 65.6% to differentiate individuals with high risk from those with low-moderate risk of falling. The other measures showed lower discriminating abilities (0.46<AUC<0.54).

## 4. DISCUSSION

This study primarily sought to establish validity of force plate measures of standing balance through identifying their concurrent validity and their ability in discriminating various known sub-groups within the sub-acute stage of stroke recovery. We hypothesized that our selected force plate measures, extracted from a 30-second standing trial, would demonstrate at least moderate concurrent validity, and would have some ability to differentiate fallers from non-fallers, or those with low-moderate versus high risk of falling. Among the measures studied here, our findings suggest that mean speed of AP-CoP has moderate concurrent validity when compared to the BBS, and WBA has the highest ability to differentiate between people with and without a history of falls, or people with low-moderate versus high risk of falling.

### 4.1. Concurrent validity

The finding of a statistically significant correlation between the mean speed of AP-CoP (but not ML-CoP) and the BBS in our study aligns with a previous study by Sawacha *et al*. (2013), which included participants with chronic stroke.^11^ In another study in chronic stroke, Frykberg *et al*. (2007) did not find a statistically significant correlation between the speed of AP-CoP and BBS; however, similar to our study, they identified a stronger correlation for speed of AP-CoP (−0.51) than for ML-CoP (−0.32).^10^ The observed similarity between the pattern of our findings and those of previous studies (i.e., stronger correlation for speed of AP-CoP than speed of ML-CoP)^10,11^ may indicate that the BBS challenges balance control more in the AP-direction than in ML-direction. Indeed, 7 out of 14 items of the BBS seem to impose a greater challenge on controlling CoP displacements along AP-direction (i.e., sitting to standing, standing unsupported, standing to sitting, transfers, standing unsupported with eyes closed, reaching forward with outstretched arm, and picking up an object from the floor), and at least one more item appears to impose an equal challenge for controlling CoP along both AP and ML-directions (i.e., standing unsupported with feet together).

In this study, we did not observe significant correlations between WBA or symmetry index and the BBS. This finding about WBA is in line with a prior study^8^ in which researchers did not find any significant correlation between WBA and other performance-based measures of mobility and balance, namely Timed Up and Go test, 10 Metre Walk test, Step test, and Functional Reach test (−0.13≤ρ≤0.14).^8^ Stroke-induced weight-bearing asymmetry arises from a combination of factors, including impaired sensory integration, visuospatial neglect, altered perception of verticality, hemiplegia, muscle weakness, and discoordination.^17,33,34^ However, performance-based balance and mobility measures, such as the BBS, may allow for achieving good scores by compensating for asymmetric weight-bearing; for example, it has been shown even in the presence of asymmetric weight-bearing (less weight on the paretic leg) or increased contribution of the non-paretic leg (higher symmetry index), individuals with sub-acute stroke could achieve a BBS score within their age-matched normative values.^35^ Thus, it is not surprising that the symmetry index, which indicates the individual-limb contributions to balance control,^16^ also does not show a significant correlation with the BBS in the present work.

Rather than suggesting the lack of concurrent validity of force plate measures, our findings imply that the BBS and force plate measures of balance measure different aspects of balance control. In addition to the points addressed in the previous paragraphs, and given the models of functioning and disability (i.e., the International Classification of Functioning, Disability, and Health, and the Nagi models), force plate measures such as speeds of CoP, weight-bearing asymmetry, and symmetry index seem to evaluate balance more at the impairment level (e.g., physiological functions of balance control systems such as muscle activity, coordination).^36^ In contrast, the BBS evaluates balance more at the functional activity level (e.g., transferring, turning, reaching).^36^

### 4.2. Ability to discriminate between sub-groups

Among the measures studied here, WBA demonstrated the best ability to differentiate sub-groups of individuals in the sub-acute stage of stroke recovery. Our WBA cut-off value for differentiating fallers from non-fallers (i.e., 47.8% of body weight on the paretic limb) agrees with a previous study that reported on the normal ranges of weight-bearing asymmetry.^37^ It has previously been shown that people with stroke who bear more weight on their non-paretic leg have more severe strokes and motor impairment of their paretic limb than those with symmetric weight-bearing.^37^ Therefore, the relationship between WBA and fall risk may simply indicate that people with more severe strokes and greater motor impairment are at increased risk of falling. In response to perturbations, most individuals with stroke prefer to take steps with their non-paretic leg,^38^ and the majority are unable to initiate reactive stepping with their non-preferred leg.^3^ However, in situations where reactive stepping by non-paretic leg is necessary to respond to a loss of balance, increased weight-bearing on the non-paretic leg may delay step initiation and increase the risk for falling.^39^ It should be noted that in our study participants were dichotomized based on their history of falls recorded during their acute-care stay, because sufficient information about participants” history of falls during inpatient rehabilitation was not available in the database used. This factor may have contributed to the negligible to low discriminative ability of the selected force plate measures, as participants” balance ability might have improved since their time in acute-care. Thus, future research using more recent fall records might be required.

### 4.3. Correlation with motor impairment and balance confidence

Our secondary objective was to examine relationships between force plate measures of standing balance and the clinical measures of paretic leg motor impairment, and perceived balance confidence. Although our results revealed only a trend of negative correlation between the paretic leg motor impairment and speeds of ML-CoP, AP-CoP and symmetry index, these findings may indicate that people with more impaired paretic leg experience more challenging balance control especially along the ML-direction. This may imply that even with a potentially greater contribution of the non-paretic limb to balance control, individuals with sub-acute stroke still experience challenges in controlling their standing balance medio-laterally.

## 5. LIMITATIONS

The BBS score is apparently influenced by individuals” compensatory strategies to control balance, rather than underlying physiological factors of impaired balance control.^35^ Thus, future studies should employ reference measures that are believed to have a better focus on the underlying aspects of balance control (e.g., reactive control, anticipatory postural adjustments, and sensory orientation mechanisms) such as the Balance Evaluation Systems Test or Mini-Balance Evaluation Systems Test.^36,40^ In addition, most of our participants were assessed during the early sub-acute stage of stroke recovery (the first 3 months), therefore our findings might not apply to people in the more chronic post-stroke stages. Furthermore, our findings might not apply to other neurological diseases, or to force plate-based assessments conducted in other test conditions (e.g., eyes closed).

## 6. CONCLUSIONS

Among the studied force plate measures of balance, mean speed of CoP along the AP direction and weight-bearing asymmetry, both calculated from a 30-second standing trial, were the measures with highest concurrent validity and discriminative ability, respectively. Use of these validated measures during the sub-acute stage of stroke recovery can provide researchers and clinicians with more clinically relevant information about the underlying mechanisms of balance impairments post-stroke. Future studies should investigate the validity of force plate measures in other stages of stroke recovery, using reference measures that mainly focus on the underlying aspects of postural balance.

## Data Availability

The participants of this study did not consent for their data to be shared publicly; therefore, supporting/raw data is not available.

## AUTHOR CONTRIBUTIONS

Raabeae Aryan: Conceptualization, Formal analysis, Methodology, Writing – original draft, Writing – review & editing. Kara K. Patterson: Methodology, Writing – review & editing, Supervision. Elizabeth L. Inness: Funding acquisition, Resources, Writing – review & editing, Data curation, Supervision. George Mochizuki: Methodology, Writing – review & editing, Supervision. Avril Mansfield: Conceptualization, Data curation, Funding acquisition, Methodology, Resources, Supervision, Writing – review & editing.

